# A Clinical Theory-Driven Deep Learning Model for Interpretable Autism Severity Prediction

**DOI:** 10.64898/2026.01.25.26344792

**Authors:** Xiyang Hu

**Affiliations:** Arizona State University

**Keywords:** Autism spectrum disorder, Deep learning, Theory-driven design, Interpretable machine learning, AI for health

## Abstract

Autism spectrum disorder (ASD) affects many children worldwide, yet clinical assessment of symptom severity remains resource-intensive and unevenly accessible. Artificial intelligence (AI) has transformative potential to support scalable and timely severity assessment from behavioral data, but existing approaches largely treat autism as a monolithic prediction target and rely on opaque models that are difficult for clinicians to interpret or trust. Moreover, prior multimodal methods typically integrate heterogeneous behavioral signals using ad hoc fusion strategies that are weakly grounded in clinical theory. We propose a clinical theory-driven deep learning model for interpretable autism severity assessment that explicitly operationalizes established clinical constructs into model design. Drawing on autism research, we represent social construct and motor construct as distinct latent components. These components are integrated through a structured cross-modal attention mechanism guided by a learnable alignment mask that encodes soft spatial correspondence priors between visual and kinematic representations. Theory-specific blocks then aggregate aligned tokens into construct embeddings, which are fused via instance-specific theory weights, yielding transparent symptom profiles aligned with clinical reasoning. Comprehensive experiments demonstrate the state-of-the-art performance of our model over existing baselines. Ablation studies validate that performance gains arise from theory-driven design choices. Analysis of the learned theory weights reveals systematic relationships between symptom profiles and severity, providing empirical support for the multidimensional structure of autism. This work demonstrates how clinical theory can be instantiated as empirically testable architectural designs in deep learning models, advancing both predictive utility and interpretability in healthcare AI systems.

## 1. Introduction

Autism spectrum disorder (ASD) represents one of the most significant neurodevelopmental conditions affecting children worldwide, with profound implications for individuals, healthcare systems, and society at large. The condition is characterized by persistent challenges in social communication and interaction, alongside restricted and repetitive patterns of behavior, interests, or activities (Guha 2014). Early and accurate identification of autism severity is critical because it directly informs the intensity and type of interventions a child receives, and research consistently demonstrates that early intervention leads to significantly improved developmental trajectories (Dawson et al. 2010). Yet the diagnostic process remains resource-intensive and inequitable: the gold-standard assessment instrument, the Autism Diagnostic Observation Schedule (ADOS, Lord et al. 2000), requires administration by extensively trained clinicians over 40 to 60 minutes of structured interaction, followed by detailed coding and interpretation. This bottleneck, with diagnostic evaluation wait times extending to 12 to 18 months in many regions, causes many children to miss critical early intervention windows, leading to long-term consequences for development and quality of life.

Recent advances in artificial intelligence (AI) offer a promising pathway to mitigate these diagnostic bottlenecks by augmenting or partially automating aspects of autism assessment. By systematically analyzing observed behavior, AI-based systems have the potential to support scalable screening, triage, and clinical decision support, particularly in resource-constrained clinical settings (Morrow et al. 2023). A growing body of work has applied machine learning and deep learning methods to autism-related tasks using video, audio, or motion data, reporting encouraging results in severity estimation and diagnostic classification (Zahan et al. 2023, Bae et al. 2025). Despite this progress, existing approaches exhibit fundamental limitations that restrict both their practical effectiveness and their readiness for integration into clinical workflows.

First, most AI-based autism assessment models treat severity as a single, undifferentiated prediction target. Behavioral signals are processed through generic architectures optimized primarily for prediction accuracy, without explicit consideration of how different symptom domains contribute to severity (Cavus et al. 2021). This stands in contrast to clinical assessment, where clinicians interpret observed behaviors through established theoretical constructs, including social communication deficits (Chevallier et al. 2012), and motor coordination difficulties (Fournier et al. 2010, Gowen and Hamilton 2013). Clinicians then integrate evidence across these domains to form severity judgments. When computational models ignore this structure, they risk capturing spurious correlations, lack the flexibility to adapt to heterogeneous symptom profiles, and provide limited insights into understanding of autism heterogeneity. Second, many existing models operate as black boxes. Although post hoc explanation techniques are sometimes applied, these explanations are often weakly connected to clinical concepts and difficult for clinicians to verify. In high-stakes healthcare contexts, lack of transparency poses a significant barrier to adoption (Rudin 2019, Durán and Jongsma 2021), as clinicians must be able to understand, evaluate, and contextualize algorithmic recommendations within their own reasoning processes. Models that produce accurate but opaque predictions provide limited value as clinical decision support tools. Third, prior multimodal approaches frequently rely on naive fusion strategies, such as feature concatenation, that do not reflect the semantic relationships between heterogeneous behavioral modalities (Zahan et al. 2023). Visual appearance and body movement, for example, capture complementary but structurally related information. Treating these modalities as interchangeable feature representations ignores their inherent asymmetries and limits a model’s capacity to integrate evidence in a manner consistent with clinical reasoning.

In this study, we propose a novel clinical theory-driven deep learning model for interpretable autism severity prediction. Rather than treating autism severity as a monolithic outcome, our model explicitly encodes two established clinical constructs, namely social communication deficits and motor control impairments, as distinct latent components. Each construct is derived from the behavioral modality that most naturally affords its measurement, reflecting clinical assessment practice. Visual representations capture appearance-based cues relevant to social communication, while kinematic representations capture movement dynamics associated with motor control. To integrate these components, we introduce a structured cross-modal attention mechanism in which visual features provide contextual queries over kinematic representations. This integration is guided by a learnable alignment mask that encodes soft spatial correspondence priors between image regions and skeletal joints, balancing domain knowledge with data-driven adaptation. After cross-modal alignment, we apply theory-specific processing blocks before projection into latent construct spaces. A Social Attention Block aggregates patch-level features through self-attention to model coordinated appearance-based patterns, while a Motor Coordination Block explicitly models bilateral structure and left-right asymmetry in joint embeddings. These blocks produce construct-aligned representations that are subsequently projected into theory-specific latent vectors. Finally, the model learns instance-specific theory weights that quantify the relative contribution of each construct to the predicted severity score. The final prediction is formed as a weighted linear combination of construct embeddings, ensuring that each theory’s contribution is directly traceable in the output. This design yields interpretable symptom profiles that decompose predictions into clinically meaningful components, providing transparency by construction rather than relying on post hoc explanation.

We evaluate the proposed model using a publicly available autism assessment dataset derived from standardized clinical interactions. Empirical results show that our model achieves state-of-the-art predictive performance while providing clinically transparent outputs. Through controlled ablation studies, we demonstrate that performance gains stem from theory-driven design choices rather than increased model complexity. Analysis of the learned theory weights further reveals systematic relationships between symptom profiles and severity, consistent with clinically observed heterogeneity across behavioral domains. These results provide empirical support for the multidimensional and heterogeneous structure of autism symptomatology in clinical theory, and demonstrate that our model not only predicts severity accurately but also recovers clinically meaningful structure that aligns with established clinical practices and theoretical expectations.

This work makes multiple contributions. First, we present the first model architecture that explicitly operationalizes established clinical theories of autism as structural components of a multimodal deep learning system, demonstrating that theory-driven design can simultaneously improve predictive performance and interpretability. Rather than relying on post-hoc explanation, our approach encodes clinical constructs directly into the model, yielding transparent, construct-level symptom representations. Second, we make a theoretical contribution by showing that clinical theories can be instantiated as empirically testable architectural designs. Through systematic ablation, we evaluate core theoretical assumptions about the relative roles of social communication and motor control in autism severity, moving beyond descriptive theory toward falsifiable computational instantiations. Third, we deliver a practical clinical decision support framework that achieves state-of-the-art predictive accuracy while producing interpretable, instance-specific symptom profiles that clinicians can directly relate to observed behavior. By enabling clinicians to inspect how different symptom domains contribute to severity estimates for each individual, the system supports trust calibration, individualized intervention planning, and integration with existing clinical workflows. Together, these contributions establish a new paradigm for autism assessment systems, in which computational models do not merely predict outcomes, but instantiate clinical theory, support clinical reasoning, and generate testable insights into symptom heterogeneity.

## 2. Related Work

### 2.1. Clinical Theories of Autism

Autism spectrum disorder is clinically defined by deficits in social communication and the presence of restricted, repetitive behaviors (Guha 2014). Recent research increasingly emphasizes autism as a multidimensional condition with motor-related differences that extend beyond the diagnostic core (Fournier et al. 2010). We focus on two theoretically grounded behavioral constructs that serve as conceptual building blocks for our architecture: social communication and motor control.

#### Social Communication Deficits

Impairments in social interaction constitute a hallmark of autism, encompassing reduced eye contact, atypical facial expressions, diminished reciprocal gestures, and unusual body posture during social exchanges (Rutter et al. 2012, Lord et al. 2000). The social motivation theory (Chevallier et al. 2012) proposes that these deficits reflect reduced intrinsic interest in social stimuli, linked to abnormalities in brain reward systems. Clinically, assessment protocols like ADOS (Lord et al. 2000, Rutter et al. 2012) heavily emphasize observation of these social-communicative behaviors, making them central targets for computational modeling. Our architecture operationalizes this construct through visual features extracted from visual representations, which preserve appearance-based cues related to body posture, spatial orientation, and movement context relevant to social communication.

#### Motor Control Impairments

Motor coordination difficulties affect 50–80% of individuals with autism (Fournier et al. 2010, Kangarani-Farahani et al. 2024), including deficits in fine motor skills, manual dexterity, balance, and bilateral coordination. Importantly, motor impairments correlate with core autism symptom severity (Gowen and Hamilton 2013, MacDonald et al. 2013), suggesting they are not merely comorbid but may reflect shared underlying neurological factors. From a computational perspective, kinematic features capturing joint coordination patterns and movement smoothness provide direct measurements of motor control. Our model explicitly represents motor control as a distinct latent channel derived from skeletal joint trajectories, enabling empirical assessment of its incremental contribution to severity prediction.

These two theoretical constructs form the conceptual foundation of our architecture, distinguishing it from atheoretical approaches that treat autism as a monolithic classification target without internal structure.

### 2.2. Computational Methods for Autism Assessment

The application of machine learning to autism assessment has evolved from traditional feature engineering approaches to modern deep learning methods, with recent work exploring multimodal integration.

#### Traditional Machine Learning with Handcrafted Features

Early computational approaches relied on expert-designed features extracted from behavioral observations, physiological sensors, or demographic data, combined with classical supervised learning algorithms (e.g., logistic regression, rule induction, decision trees, support vector machines, random forests) (Wall et al. 2012, Duda et al. 2016). While these methods may benefit from interpretability and data efficiency, their performance is bounded by the quality and completeness of manually engineered features. They also struggle to capture complex, hierarchical patterns in high-dimensional behavioral data such as video sequences. Our experiments include traditional ML baselines to establish whether deep learning provides tangible improvements over expert feature engineering.

#### Single-Modality Deep Learning

Recent work applies deep neural networks to autism-related prediction tasks using individual data modalities. Convolutional neural networks (CNNs) have been used for facial expression analysis, while recurrent neural networks (RNNs) and graph convolutional networks (GCNs) have been applied to time-series physiological data and skeletal motion sequences. Pose-based models capture motor patterns (Zhang et al. 2021), video-based methods detect affective and social cues (Hashemi et al. 2018), and acoustic models extract speech prosody (Bone et al. 2016). These approaches achieve impressive performance on specific modalities but ignore the complementary information available from multimodal observations. Our single-modality baselines test whether multimodal integration is helpful for this task.

#### Multimodal Learning for Autism

A growing body of work recognizes that autism manifests through multiple observable channels, motivating multimodal approaches. Tariq et al. (2018) used smartphone videos and behavioral ratings for autism classification. More recent methods align text, audio, and video cues for early screening (Zhu et al. 2023, Bae et al. 2025). Zahan et al. (2023) proposed a state-of-the-art multimodal method combining visual and kinematic data for autism detection, demonstrating performance improvements over single-modality approaches. However, for all these methods, behavioral signals from different modalities are typically fused through feature concatenation or late-stage aggregation, without explicit modeling of the semantic relationships between modalities or aligning model structure with clinical constructs. As a result, these models provide limited insight into how different symptom domains contribute to severity estimates and offer little support for understanding symptom heterogeneity. Our work advances these by introducing cross-attention for semantic correspondence learning and theory-guided architecture design for interpretable, clinically grounded integration.

### 2.3. Theory-Driven and Interpretable AI in Healthcare

Our work also contributes to the call for theory-driven design of AI algorithms, particularly in high-stakes domains like healthcare where interpretability and trust are paramount.

#### Interpretable Machine Learning for Clinical Decision Support

The black box nature of deep learning poses challenges for clinical adoption, where predictions must be explainable to support decision-making and ensure accountability (Durán and Jongsma 2021, Ahmed et al. 2023). Interpretable ML methods, including attention visualization, saliency maps, concept-based explanations, and inherently interpretable architectures, aim to open these black boxes (Rudin et al. 2022). However, many interpretability techniques are completely *post-hoc* (explaining existing models after training) rather than *by-design* (building interpretability into architecture), which have concerning reliability limitations (Rudin 2019). Our approach embodies interpretability-by-design: theoretical constructs are explicit latent channels, and learnable theory weights provide instance-specific explanations in terms of clinically meaningful symptom dimensions. These ensure predictions are not only accurate but also *intelligible* to clinicians and *grounded* in established theories.

#### Theory-Driven Model Design for Healthcare AI

Recent research has increasingly incorporated medical and behavioral theory into the design of healthcare algorithms. By organizing model architectures around medical meaningful constructs, these approaches seek to improve both predictive performance and interpretability. For instance, Xie et al. (2025) propose a knowledge-guided neural topic model that incorporates medically grounded seed terms to predict the mental health effects of online video content. Li et al. (2025) introduce a large language model multi-agent framework for drug discovery that is guided by organizational theories of cognitive labor and iterative learning, supporting coordinated agent collaboration with biomedical knowledge grounding. Zhou et al. (2023) develop a multi-armed bandit algorithm for personalized health intervention recommendation, where diversity promotion is informed by behavioral theory. Morid and Sheng (2025) present a channel-wise deep learning model for patient cost prediction that structures inputs by distinct categories of medical codes, addressing both data heterogeneity and sociotechnical fairness. Lin et al. (2017) propose a multitask learning framework for chronic care risk profiling, where each task corresponds to a specific adverse outcome and shared structure supports joint clinical inference. Our model follows this line of work by aligning modality-specific channels with behavioral constructs derived from autism theory.

## 3. Methodology

### 3.1. Task Definition

We study the problem of predicting autism severity from *temporal skeleton sequences*, which represent body joint positions over time. In many clinical and research settings, raw video recordings of behavioral assessments are not available for downstream modeling. Due to privacy regulations governing sensitive recordings of children, including HIPAA and GDPR, video data are often not released in their original form and are instead transformed into privacy-reduced representations suitable for analysis (Ahmed et al. 2023). Among these, skeleton sequences are widely used and commonly distributed in autism datasets, making skeleton-based modeling a prevalent practical setting (Billing et al. 2020). In addition to their availability, skeleton representations remove identifiable visual information such as facial appearance, clothing, and scene context while preserving kinematic structure relevant to behavior (Cao et al. 2019). This combination of practical accessibility and reduced privacy risk makes temporal skeleton sequences a common representation for autism assessment.

Formally, let *y* denote the ground-truth ADOS total score for an individual, where higher scores indicate greater symptom severity (Lord et al. 2000, Gotham et al. 2009). The ADOS total score aggregates assessments across multiple behavioral domains, including social affect, restricted and repetitive behaviors, and overall impression, making it a clinically validated summary measure of autism severity (Rutter et al. 2012). Our prediction task is formulated as a regression problem: given a skeleton temporal sequence χ∈ ℝ^*C*×*T*×*J*^capturing an individual’s movements during a standardized assessment session (where *C* denotes spatial coordinates, *T* denotes temporal frames, and *J* denotes body joints), predict the ADOS total score *ŷ* ≈ *y*.

Following prior work (Zahan et al. 2023), we derive two complementary representations from the same underlying skeletal sequence. First, we retain χ in its native form as *kinematic data*, which captures joint-level movement dynamics suitable for modeling motor control. Second, we transform χ into a *SKEPXEL pseudoimage* ℐ via a skeleton-to-pixel projection (Liu et al. 2019), which encodes spatiotemporal joint trajectories into a two-dimensional visual format. Although both representations originate from the same skeletal signal, they induce distinct structural organizations and learning biases. The native skeleton representation preserves explicit anatomical topology and temporal continuity, supporting graph-based modeling of joint connectivity and coordinated motion. In contrast, the SKEPXEL transformation reorganizes trajectories into a spatially arranged grid of superpixels, enabling patch-based processing through vision Transformer architectures that emphasize global spatial relationships and configurational patterns. As a result, the two encodings induce different representational geometries and inductive biases prior to shared latent projection: one structured by graph connectivity and temporal convolution, and the other structured by two-dimensional spatial embedding and self-attention. This setting can be understood as representation-level multimodality, where heterogeneity arises from structural encoding and computational inductive bias rather than distinct sensing devices. The heterogeneity arises from structural encoding and computational inductive bias rather than from different sensing devices. Under privacy constraints that preclude release of raw video data, this strategy enables heterogeneous feature extraction from a single privacy-preserving source. Empirically, as shown in § 5, combining these structurally distinct encodings improves predictive performance relative to either representation alone.

### 3.2. Clinical Theory-Driven Design Rationale

Our model design is informed by clinical theory and practice. We summarize our high-level design rationale:

First, construct-level representation. Clinical assessment of autism severity distinguishes among multiple symptom domains, including social communication and motor coordination. Rather than treating behavioral data as undifferentiated input, we adopt a theory-driven approach that explicitly operationalizes *two* clinical constructs, namely *social communication* and *motor control*, as distinct latent channels within the model architecture. This decomposition mirrors clinical reasoning, enables interpretable predictions by exposing each symptom dimension’s contribution, and supports empirical analysis of which domains are most predictive.

Second, modality-construct alignment. Different behavioral domains are most naturally observed through different modalities. Appearance-based cues are central to assessing social communication behaviors, while joint kinematics provide direct evidence of motor control. Accordingly, social communication is modeled from visual features derived from SKEPXEL pseudo-images that capture holistic body configurations, whereas motor control is modeled from skeleton joint trajectories. This alignment grounds each construct in the observational channel that best supports its measurement in clinically meaningful ways.

Third, instance-specific interpretability under heterogeneity. Autism is characterized by substantial inter-individual variability, with different individuals exhibiting different patterns of strengths and impairments across symptom domains. To reflect this heterogeneity, the model learns instance-specific weights over the construct-level components rather than relying on fixed fusion rules. These learned weights indicate the relative contribution of social and motor dimensions to each individual’s predicted severity and can be interpreted as personalized symptom profiles, mirroring how clinicians conceptualize autism heterogeneity.

Figure 1 illustrates our model architecture, which operationalizes the above principles through a modular separation of feature extraction and theory-guided integration. Visual and kinematic inputs are processed by frozen pretrained encoders, after which cross-attention aligns modalities. Theory-specific blocks (Social Attention Block for patches, Motor Coordination Block for joints) then aggregate the aligned tokens in a theory-consistent way, and the resulting vectors are projected into two construct-specific latent channels corresponding to social communication and motor control. These channels are integrated through learnable, instance-specific weighting for severity prediction. We employ cross-modal attention mechanisms to allow effective information exchange between modalities, with a learnable alignment mask that encodes soft priors over spatial correspondence between image patches and skeleton joints, such as stronger coupling between head-region patches and head or neck joints. We present each component of the architecture in detail below.

**Figure 1.**
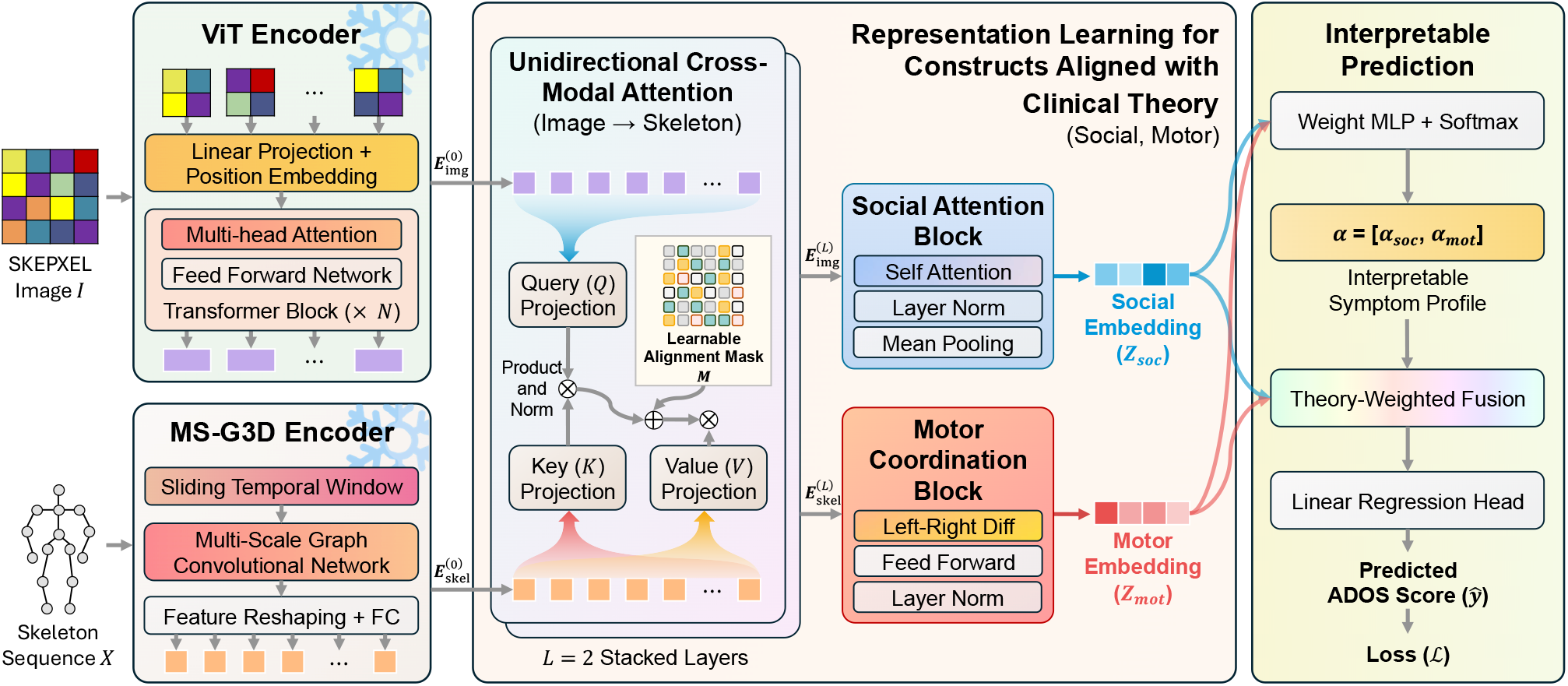
Overview of the proposed theory-guided model architecture for autism severity prediction. A SKEPXEL pseudo-image and a skeleton sequence are encoded by frozen pretrained backbones (ViT and MS-G3D) into patch and joint tokens, which are aligned via unidirectional cross-attention from image patches to skeleton joints. A learnable alignment mask biases attention toward plausible joint-to-patch correspondences. The aligned representations are then processed by theory-specific blocks (Social Attention Block over patches, Motor Coordination Block over joints) and projected into two theory-aligned latent channels (social communication, motor control), combined using instance-specific theory weights, and passed through a linear regression head to produce the final severity estimate.

### 3.3. Multimodal Feature Extraction: Frozen Domain-General Encoders

Given a multimodal input consisting of a skeleton sequence χ and its corresponding visual representation ℐ, our model first performs feature extraction using frozen pretrained encoders. Each modality is transformed into high-dimensional embeddings that capture kinematic and visual information at appropriate temporal and spatial resolutions. These embeddings are treated as fixed, semantically informative representations and are subsequently aligned and integrated by task-specific, theory-driven modules described in later subsections.

This design leverages large-scale pretrained models to provide robust and transferable feature extractors for graph-structured motion data and images, while concentrating learning capacity on the autism assessment-specific problem of interpreting and integrating these features in accordance with established clinical theories. At the same time, freezing the pretrained encoders sharply limits the number of trainable parameters, reducing the risk of overfitting on small clinical datasets. Such datasets are typically orders of magnitude smaller than those used for pretraining vision and action recognition models, making end-to-end fine-tuning prone to memorization rather than generalization.

#### 3.3.1. Kinematic Encoder: MS-G3D Graph Convolutional Network for Skeleton Sequences

To extract kinematic features from the skeleton sequence χ, we employ a Multi-Scale Graph Convolutional Network with 3D convolutions (MS-G3D) (Liu et al. 2020), a state-of-the-art architecture specifically designed for skeleton-based action recognition. The MS-G3D model represents skeleton sequences as spatiotemporal graphs, where nodes correspond to body joints and edges encode physical connectivity (bone structure) and temporal links across frames.

##### Graph Representation

The skeleton graph is defined by a binary adjacency matrix that encodes the skeletal topology (e.g., the wrist is connected to the elbow, the elbow to the shoulder, etc.). This structural prior is essential for learning physically meaningful movement patterns, as it constrains the model to respect anatomical constraints when aggregating information across joints.

##### Multi-Scale Spatiotemporal Convolutions

The MS-G3D architecture applies graph convolutions at multiple spatial scales (capturing both local joint relationships and longer-range kinematic chains) and temporal scales (capturing both short-term motion dynamics and longer-term movement trends). The multi-scale design is motivated by the observation that different autism-related motor behaviors manifest at different spatiotemporal scales: for example, fine motor coordination may be reflected in localized, rapid joint movements, while other body movement asymmetry may be evident in longer-range, slower bilateral patterns.

After processing the input skeleton sequence χ through multiple MS-G3D layers, the model produces a feature tensor with dimensions (*N, d, T* ^′^, *J*), where *N* is the batch size, *d* is the learned feature channel dimension, *T* ^′^ is the temporally downsampled time dimension, and *J* is the number of joints. To obtain joint-level embeddings suitable for cross-modal alignment, we perform temporal average pooling: 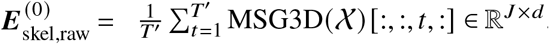 . This yields a set of *J* joint embeddings, each of dimensionality *d*, that encode the kinematic patterns associated with each body joint over the entire observed sequence.

##### Projection

We apply a trainable linear projection independently to each joint token: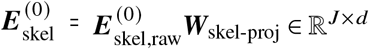 . This projection functions as an alignment and adaptation layer, mapping pretrained kinematic representations into the shared latent space used by subsequent multimodal fusion modules.

From a theoretical perspective, the frozen MS-G3D encoder captures *kinematic behavioral features* that are directly relevant to motor control assessment. The joint-level representations encode movement dynamics, coordination patterns, joint angle trajectories, and temporal rhythms that reflect underlying motor capabilities and locomotion characteristics.

#### 3.3.2. Visual Encoder: Vision Transformer for SKEPXEL Images

To extract visual features from the SKEPXEL pseudo-images ℐ, we employ a Vision Transformer (ViT) architecture (Dosovitskiy et al. 2021). The ViT model processes images by partitioning them into a regular grid of non-overlapping patches, embedding each patch via a learned linear projection, and then applying a stack of Transformer encoder layers to capture global relationships among patches through self-attention.

##### Patch Tokenization

Given an input SKEPXEL image ℐ ∈ ℝ^*H*×*W*×3^, the ViT divides the image into a 14 × 14 grid, yielding *P* = 196 patches of size 16 × 16 pixels each. Each patch is flattened and linearly projected to produce a *D*_ViT_ = 768-dimensional embedding. After adding learned positional encodings (to preserve spatial structure) and passing through the ViT’s Transformer encoder stack, the model outputs a set of patch-level embeddings: 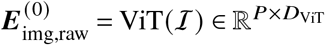 .

##### Projection

To support direct cross-modal interaction with joint-level kinematic tokens, we apply a learned linear projection to each visual patch embedding, mapping the ViT output from *D*_ViT_ into the shared latent space of dimension 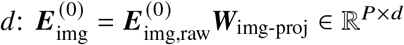. This projection aligns appearance-based visual features with the kinematic embedding space, enabling meaningful comparison, correspondence modeling, and fusion between visual patches and body joints in downstream multimodal attention mechanisms. The projection layer is trainable, allowing adaptation of pretrained ViT features to the SKEPXEL representation and the target behavioral assessment task.

From a theoretical perspective, the frozen ViT encoder captures *appearance-based behavioral cues* that are relevant to social communication assessment. The patch-level representations encode visual patterns such as body posture configurations, spatial orientations, head positions, and overall movement contexts. These visual features complement the more explicitly kinematic information provided by the skeleton encoder, enabling the model to integrate complementary perspectives on the same behavioral episode.

### 3.4. Cross-Modal Alignment via Unidirectional Attention: Image-to-Skeleton Information Flow

A critical challenge in multimodal fusion for autism assessment is aligning and integrating information from heterogeneous modalities, visual (patch-based image features) and kinematic (joint-based skeleton features), that represent complementary perspectives on the same behavioral episode. Our architecture addresses this through a unidirectional cross-attention mechanism in which *image patches query skeleton joints*, enabling visual contextual features to augment kinematic representations in a structured and interpretable manner.

#### 3.4.1. Theoretical Motivation for Unidirectional Image-to-Skeleton Attention

We design the attention flow from image to skeleton (rather than bidirectional or skeleton-to-image) based on the following clinical considerations:

First, visual appearance cues, such as overall body posture, spatial orientation relative to interaction partners, head position, and configurational patterns, provide rich contextual information that is highly relevant for assessing social communication deficits, a core dimension of autism symptomatology. By allowing image patches to query skeleton joints, the model can effectively ask: “Given the visual context, how should we interpret the kinematic patterns of specific joints?” This query direction aligns with clinical reasoning process (Guha 2014), where contextual visual cues inform the interpretation of specific movements.

Second, the SKEPXEL representation inherently encodes spatiotemporal patterns at a more global level compared to individual joint trajectories. Image patches capture not only individual joint configurations but also their spatial relationships, movement trajectories over time, and overall movement “gestalt.” Directing attention from this richer, more contextual modality (image) to the more local, anatomically structured modality (skeleton) allows the model to use global patterns to guide integration of local kinematic features.

Third, from a regularization perspective, unidirectional attention imposes a simpler, more constrained integration structure compared to bidirectional cross-attention, which may help prevent overfitting on small clinical datasets. By restricting information flow to a single direction, we reduce the number of trainable parameters and the complexity of cross-modal interactions, potentially improving generalization. Our experimental results (reported in §5.5) empirically validate this design choice, demonstrating that unidirectional image-to-skeleton attention outperforms both bidirectional and reverse-direction alternatives.

#### 3.4.2. Learnable Alignment Mask: Encoding Spatial Correspondence Priors

A key innovation in our cross-attention design is the introduction of a learnable alignment mask ***M*** ∈ ℝ^*J*×*P*^ that acts as an additive bias on attention logits. This mask encodes soft priors over the correspondence between skeleton joints and image patches, reflecting certain joints naturally attend more strongly to specific image regions. Consider, for example, the correspondence between the head joint and the upper-center region of the image, or between hand joints and patches near the extremities. While these correspondences are not perfectly deterministic (they depend on body pose, camera viewpoint, and movement dynamics), they provide useful structural priors that guide the attention mechanism toward anatomically and spatially coherent alignments.

##### Learning vs. Imposing Correspondence

Rather than manually specifying these correspondences (which would require labor-intensive annotation and may not generalize across different pose configurations), we make the alignment mask ***M*** a *learnable parameter* of the model. The mask is initialized with small random values and updated via gradient descent during training, allowing the model to discover the most predictive correspondence patterns directly from the data. This design balances domain knowledge (spatial structure matters) with flexibility (let data determine the specific correspondences).

#### 3.4.3. Multi-Layer Cross-Attention Architecture

Our cross-modal integration module consists of *L* = 2 stacked cross-attention layers, enabling hierarchical refinement of multimodal representations. Let 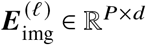 and 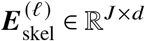 denote the patch and joint embeddings at layer *𝓁* ∈ {0, 1, …, *L* − 1}, where 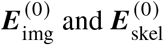 are the outputs from the frozen ViT and MS-G3D encoders as defined above.

##### Cross-Attention from Image to Skeleton

At each layer *𝓁*, we compute cross-attention in which image patches query skeleton joints. First, we form query, key, and value projections: 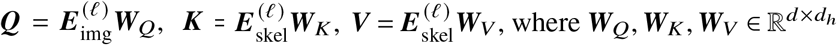 are learned projection matrices, and *d*_*h*_ is the dimensionality per attention head. We use multi-head attention with multiple parallel heads to capture diverse alignment patterns.

Next, we compute the attention weights with the learnable alignment mask:

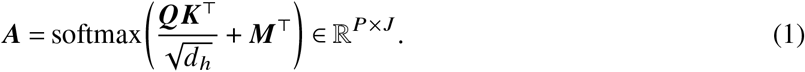

The attention matrix ***A*** encodes, for each image patch, a distribution over skeleton joints indicating which joints are most relevant for interpreting that patch. The alignment mask ***M***^⊤^ biases these weights toward spatially and anatomically coherent correspondences.

Finally, we aggregate joint features to update patch embeddings: 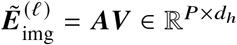. After multihead concatenation and output projection, the updated patch embeddings 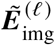 are combined with residual connections and layer normalization (following standard Transformer design) to produce 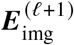.

##### Self-Attention and Feed-Forward Layers

In addition to cross-attention, each layer also includes: (1) *Modality-specific self-attention* over image patches (allowing patches to contextualize each other within the visual modality) and over skeleton joints (allowing joints to contextualize each other within the kinematic modality). These self-attention operations preserve and refine within-modality coherence. (2) *Position-wise feed-forward networks* applied independently to each token (patch or joint), providing non-linear transformation capacity. (3) *Residual connections and layer normalization* around each sub-layer, ensuring stable gradient flow and facilitating deep architecture training.

Importantly, there is *no reverse cross-attention* from skeleton joints to image patches in this architecture. This unidirectional design constrains the information flow to align with our theoretical hypothesis about the role of visual context in augmenting kinematic interpretation. The empirical validation of this design choice, compared to bidirectional and reverse-direction alternatives, is presented in §5.5.

After *L* = 2 layers of cross-attention processing, we obtain final patch embeddings 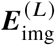 and joint embeddings 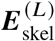 that are enriched with cross-modal information, ready for theory-guided fusion.

### 3.5. Theory-Guided Latent Decomposition and Interpretable Fusion

After the cross-attention layers align and integrate visual and kinematic modalities, the next stage of our architecture operationalizes the two theoretical constructs, namely social communication and motor control, as explicit latent channels. Rather than simply pooling cross-attended tokens and then projecting, we apply *theory-specific processing blocks* that aggregate token-level representations in a theory-consistent way before projection into latent channels. This theory-driven decomposition serves both predictive and interpretive functions: it structures the feature space according to clinically grounded dimensions, and enables instance-specific assessment of which symptom domains are most relevant for each individual’s predicted severity.

#### 3.5.1. Theory-Specific Processing Blocks and Construct-Level Representation

Instead of treating token-level embeddings as undifferentiated features, these theory-specific processing blocks aggregate modality-specific information in a manner consistent with how each construct is conceptualized and assessed in clinical practice. In this sense, construct-modality alignment is encoded directly in the architecture before any latent projection is performed.

##### Social Attention Block

Social communication deficits in autism are primarily expressed through appearance-based behavioral cues, including body posture during interaction, spatial orientation toward an interaction partner, head position, and configurational patterns that signal engagement or withdrawal. Clinically, these cues are interpreted holistically: the meaning of any single region (e.g., head orientation) depends on its relation to other regions (e.g., torso alignment or gesture configuration).

To reflect this integrative structure, we apply a *Social Attention Block* to the patch-level embeddings 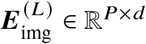. The block consists of multi-head self-attention over the *P* patches, followed by a residual connection and layer normalization, and finally mean pooling across patches:

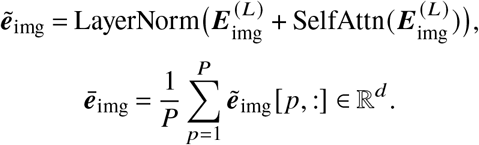

The self-attention mechanism enables the model to learn which spatial regions co-vary in a socially meaningful way, capturing coordinated posture and configurational patterns that are central to social evaluation. The resulting vector 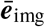 is therefore not a generic visual summary, but a construct-informed representation structured around appearance-based evidence relevant to social communication.

##### Motor Coordination Block

Motor coordination deficits in autism are most directly observable through joint kinematics, including coordination across multiple joints, movement smoothness, execution precision, and bilateral symmetry. Empirical research documents increased left-right asymmetry and coordination differences in individuals with autism (Fournier et al. 2010, Gowen and Hamilton 2013). Clinically, motor characteristics are assessed by examining how joints work together across the body, rather than by inspecting isolated joint trajectories.

To encode this structure, we apply a *Motor Coordination Block* to the joint-level embeddings 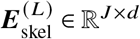. The block partitions joints into left and right body groups and computes mean feature vectors 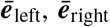 and a global mean 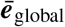. An asymmetry representation is then formed as

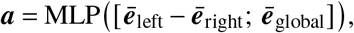

and the final output is

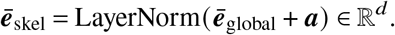

By explicitly modeling bilateral structure and asymmetry, the block biases the representation toward clinically relevant motor coordination patterns. The resulting vector 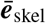 thus encodes kinematic evidence in a manner aligned with how motor control differences are theoretically understood and behaviorally assessed.

##### Projection into Latent Construct Space

The construct-informed vectors are subsequently mapped into a shared latent space through two lightweight projection functions:

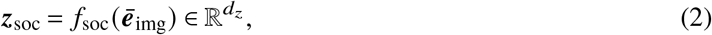

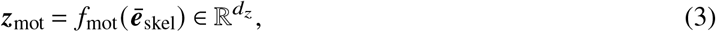

where each *f*_·_ (·) is a small multi-layer perceptron with layer normalization and dropout. These functions serve to adapt the construct-level representations to the shared latent space used for downstream fusion, without introducing additional theoretical structure.

This architectural separation mirrors clinical observation practice: social communication is primarily inferred from appearance-based interaction patterns, whereas motor characteristics are inferred from movement execution and coordination. By embedding this alignment into the representation learning stage, the model encodes theoretical structure directly into its intermediate computations.

#### 3.5.2. Learnable Theory Weights: Interpretable Instance-Specific Fusion

A critical innovation in our design is the computation of *instance-specific, interpretable theory weights* that indicate the relative contribution of each theoretical construct to the predicted severity for each individual. This design acknowledges the well-documented heterogeneity in autism symptom profiles: different individuals exhibit different patterns of strengths and deficits across social and motor domains. Rather than applying a fixed, population-level fusion rule, our model learns to adaptively weight the two latent channels based on the specific behavioral patterns observed in each case.

##### Weight Computation

We concatenate the two latent vectors to form a joint representation: 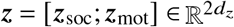. From this concatenated vector, we compute theory weight logits via a small MLP *g*(·): ***u*** = *g*(***z***) ∈ ℝ^2^, where ***u*** = [*u*_soc_, *u*_mot_]⊤ are raw, unnormalized scores for each theory channel.

To obtain interpretable, normalized weights, we apply softmax normalization with a temperature τ:

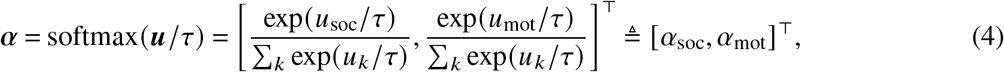

where *τ* is a hyperparameter that controls the smoothness of the weight distribution. A higher *τ* produces more uniform weights, while lower τ produces more peaked, selective weights. The resulting weight vector *α* satisfies *α*_*i*_ ≥ 0 and Σ_*i*_ *α*_*i*_ = 1, making it directly interpretable as a probability distribution over theory channels. For example, if a particular individual’s behavioral patterns yield *α*_mot_ = 0.6, *α*_soc_ = 0.4, this indicates that motor coordination deficits are the stronger predictor of severity for this individual, with social communication playing a secondary role. Such interpretable decompositions can support clinical hypothesis generation, personalized assessment, and identification of subgroups with distinct symptom profiles.

#### 3.5.3. Final Fusion and Severity Prediction

The final prediction uses a *pure weighted sum* of the theory-aligned latent channels, without any nonlinear interaction term. This design maximizes interpretability: the fused representation is a convex combination of the two construct embeddings, and the prediction is a linear function of that combination. Formally,

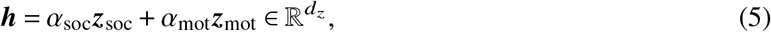

where *α* are the instance-specific theory weights defined above (normalized via softmax). The severity prediction is then

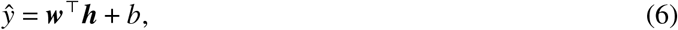

where 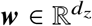 and *b* ∈ ℝ are learned parameters of a linear regression head. By avoiding an MLP or interaction term on the concatenated latents, the mapping from theory channels to prediction remains fully linear: each theory contributes in proportion to its weight, and the regression coefficients ***w*** apply uniformly to the fused vector. This yields the most interpretable fusion at the cost of limiting the model’s capacity to capture nonlinear interactions among symptom dimensions; we adopt it to prioritize transparency and clinical interpretability.

Our training objective is to minimize the mean squared error between predicted and ground-truth ADOS total scores: 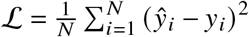, which is a standard objective for continuous regression tasks. *N* is the number of training samples, *ŷ*_*i*_ is the model’s predicted severity for sample *i*, and *y*_*i*_ is the ground-truth ADOS total score.

## 4. Experiment Setup

### 4.1. Dataset

We evaluate our model using a dataset collected in the context of structured clinical interactions with children diagnosed with autism spectrum disorder (Billing et al. 2020). The dataset was designed to support data-driven analysis of autism-related behaviors under strict privacy constraints. As raw video and audio recordings of children cannot be released, the dataset provides skeletal joint trajectories derived from multimodal sensing during standardized assessment sessions. The dataset comprises 3,121 skeletal sequences collected during standardized clinical assessment sessions using the Autism Diagnostic Observation Schedule (ADOS) for children between the ages of 3 and 6. Each sequence is associated with an ADOS total score assigned by a trained clinician.

Given the limited size of the dataset (common in clinical research), we employ *k-fold cross-validation* to obtain robust estimates of model performance and reduce the impact of dataset-specific variability. The full dataset is partitioned into *k* = 10 folds, and the training and testing procedure is repeated *k* times. Final performance metrics are reported as the mean across all folds.

### 4.2. Baseline Methods

To contextualize the performance of our proposed theory-driven multimodal architecture, we compare against multiple categories of baseline approaches:

#### Traditional Machine Learning with Handcrafted Features

These baselines represent feature-engineering approaches common in clinical informatics. We extract a comprehensive set of handcrafted features from skeleton sequences, including: joint angle statistics, movement velocity and acceleration distributions, bilateral symmetry indices (left-right coordination), and postural stability measures (center-of-mass variance), and motor and locomotion-related parameters. Three classical regressors are trained on these features: (1) XGBoost, (2) Random Forest, and (3) Support Vector Regression (SVR).

#### Single-Modality Deep Learning Approaches

These baselines process either visual or kinematic data in isolation, testing whether multimodal integration is necessary. (4) *ViT-Only*: A vision transformer pretrained on ImageNet processes SKEPXEL pseudo-images, followed by a regression head. (5) *MS-G3D-Only*: A Multi-Scale Graph Convolutional Network (Liu et al. 2020) pretrained on skeleton-based action recognition processes skeleton sequences, followed by a regression head.

#### Multimodal Baselines

(6) *Established Multimodal ADOS Baseline*: This baseline concatenates features from frozen ViT and MS-G3D encoders (the same backbones used in our approach) and passes the concatenated vector through a simple feedforward network for regression. This method represents the current state-of-the-art autism analysis approach based on skeletal joint trajectories, proposed by Zahan et al. (2023). Comparing against this state-of-the-art baseline isolates the contribution of our architectural innovations, including cross-attention and theory-guided fusion, beyond simply using multimodal data, demonstrating whether sophisticated integration mechanisms provide value over straightforward feature concatenation. (7) *Medical Vision-Language Foundation Model (MedGemma)*: Recent medical vision-language foundation models have shown strong performance across a range of clinical perception and reasoning tasks. To evaluate whether such general-purpose medical VLMs can serve as competitive alternatives to domain-specific multimodal architectures, we include MedGemma (Sellergren et al. 2025). In this setup, SKEPXEL pseudo-images are paired with a templated textual prompt describing the assessment task, and the MedGemma vision-language encoder is used to produce an ADOS prediction.

#### Ablation Variants

To isolate specific design choices in our architecture, we compare against ablated versions of our full model: (8–9) *Theory Component Ablations*: Variants with social component, or motor component removed, testing each component’s contribution. (10–12) *Fusion Design Ablations*: Variants with per theory prediction, mixture of experts (MOE), or gated fusion, testing fusion mechanism choices. (13–14) *Alignment Mask Ablations*: Variants with no alignment mask, or fixed alignment mask, testing the contribution of learnable alignment mask design. (15) *Bidirectional Cross-Attention*: Enables both skeleton-to-image and image-to-skeleton attention, testing whether unidirectional flow is superior. (16) *Skeleton-to-Image Cross-Attention*: Reverses the query direction, testing attention directionality.

### 4.3. Evaluation Metrics

Consistent with clinical assessment requirements and machine learning best practices, we evaluate model performance using three complementary metrics that capture distinct facets of performance:

#### Mean Absolute Error (MAE)

MAE measures the average magnitude of prediction errors in ADOS total scores, providing a direct interpretation in clinically meaningful units. Lower MAE indicates more accurate predictions.

#### Pearson Correlation Coefficient

Pearson correlation measures the strength and direction of the linear relationship between predicted and actual severity scores. Pearson correlation ranges from -1 to 1, with values closer to 1 indicating stronger positive linear association.

#### Quadratic Weighted Kappa (QWK)

QWK measures agreement between predicted and actual severity categories when continuous scores are binned into ordinal classes, with quadratic penalties for larger disagreements. For autism assessment, following the standardized ADOS calibrated severity score methodology (Gotham et al. 2009), we bin predicted and actual ADOS total scores into three clinically meaningful diagnostic categories based on age and evaluation module: *Non-Spectrum* (NS), *Autism Spectrum Disorder* (ASD), and *Autistic* (AUT, representing the most severe presentation). This classification accounts for developmental differences across age groups and assessment contexts, providing a more clinically valid categorization than arbitrary score cutoffs. QWK ranges from -1 to 1, with 1 indicating perfect agreement and 0 indicating agreement no better than chance. This metric is particularly relevant for clinical practice, where distinguishing between non-spectrum, ASD, and autistic presentations is more critical than precise score prediction, as these categories directly inform diagnostic conclusions and intervention intensity levels.

The three metrics jointly provide a comprehensive evaluation. MAE quantifies absolute accuracy (critical for score-based diagnosis), Pearson correlation captures rank ordering (critical for population stratification and resource allocation), and QWK measures categorical agreement (critical for severity-based intervention planning). A model that performs well across all three metrics demonstrates robust predictive utility suitable for clinical decision support systems. Reporting all three metrics also guards against overfitting to a single objective and provides multiple perspectives on model performance aligned with different clinical use cases.

## 5. Results

### 5.1. Comparison with Baseline Methods

We begin by establishing whether our theory-driven model architecture delivers tangible performance improvements over baseline approaches. We compare our model against three categories of baselines representing the current state of practice: traditional machine learning methods based on handcrafted kinematic features, single-modality deep learning models that process either visual or skeletal data in isolation, established multimodal models for autism severity prediction, and a large medical vision-language foundation model (MedGemma) that provides implicit multimodal alignment through large-scale pretraining. Table 1 summarizes the comparative results.

**Table 1.**
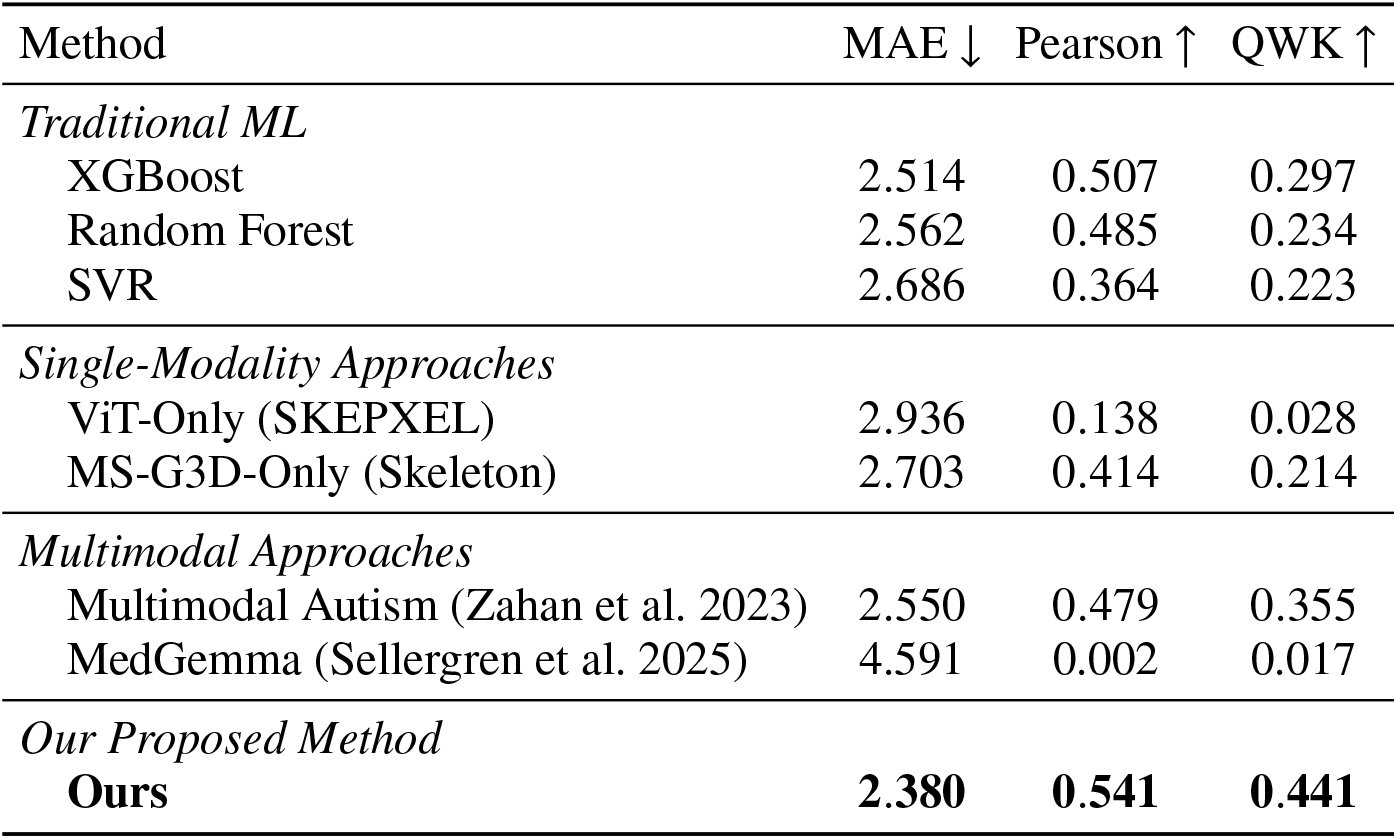
Performance Comparison with Baseline Methods.

Our proposed model demonstrates substantial improvements over all baseline methods across all three evaluation metrics. When compared to traditional machine learning approaches using handcrafted features, our method achieves consistent gains: MAE reductions of 5.3%, 7.1%, and 11.4% relative to XGBoost, Random Forest, and SVR respectively. More notably, our model attains a QWK of 0.441 compared to XGBoost’s 0.297, corresponding to a 48.5% relative improvement. This indicates substantially stronger agreement with clinically meaningful severity categories, which is particularly relevant for downstream screening and intervention planning. These gains suggest that pretrained representation learning captures hierarchical and subtle behavioral patterns that are difficult to encode through manually designed features, and that theory-guided fusion in learned latent spaces enables more nuanced integration than is possible with fixed feature sets.

Relative to single-modality approaches, our method achieves 18.9% and 11.9% MAE reductions compared to ViT-Only and MS-G3D-Only respectively, with Pearson correlation improvements of 292.0% and 30.7%. The particularly weak correlation of the ViT-Only model indicates that visual appearance cues alone provide insufficient signal for reliable severity estimation. Skeleton-based modeling performs better, reflecting the relevance of motor and kinematic patterns, but still underperforms the multimodal setting. These results support the view that autism-related behavioral severity reflects a combination of observable social cues and underlying movement dynamics, neither of which is adequately captured in isolation.

The comparison with the established multimodal autism assessment model of Zahan et al. (2023) is especially informative, which concatenates ViT and MS-G3D features without cross-attention or theory-guided integration. While this naive multimodal approach (MAE=2.550, Pearson=0.479, QWK=0.355) outperforms individual unimodal methods, it remains inferior to our theory-driven architecture. Our model achieves a 6.7% reduction in MAE (2.380 vs. 2.550), an 12.9% improvement in Pearson correlation (0.541 vs. 0.479), and a 24.2% improvement in QWK (0.441 vs. 0.355). These gains demonstrate that performance improvements arise not merely from combining modalities, but from how they are integrated. Cross-attention enables adaptive, semantically aligned interaction between modalities, while theory-guided fusion imposes clinically grounded structure that mitigates reliance on spurious correlations in high-dimensional concatenated feature spaces.

Finally, we observe that the large medical vision-language foundation model MedGemma performs poorly on this task, with near-zero correlation and substantially higher error across all metrics. This result highlights an important limitation of general-purpose medical VLMs for fine-grained behavioral assessment. Although MedGemma benefits from large-scale multimodal pretraining, its representations are optimized for broad clinical perception and reporting tasks rather than subtle temporal, kinematic, and interactional patterns that characterize autism severity. The large performance gap suggests that a powerful medical generative AI model alone is insufficient for this domain, and that task-specific architectural design and domain theory remain critical for accurate autism behavioral modeling.

### 5.2. Contribution of Theory-Guided Components

Having established the model’s overall performance advantage, we next examine whether this utility is achieved in a *theory-consistent* manner. Specifically, do the theoretical constructs we have operationalized (social communication deficits and motor control impairments) provide measurable incremental value beyond generic multimodal learning?

We conduct systematic ablation studies, removing each theoretical component while holding the architecture constant. Table 2 presents performance when each theory module is ablated.

**Table 2.** Contribution Analysis of Theory-Guided Components.

| Configuration | MAE ↓ | Pearson ↑ | QWK ↑ |
| --- | --- | --- | --- |
| Without Social Component | 2.472 | 0.507 | 0.407 |
| Without Motor Component | 2.508 | 0.484 | 0.387 |
| **Ours (Social & Motor)** | **2.380** | **0.541** | **0.441** |

All two theoretical components contribute meaningfully to predictive performance, with each removal causing measurable degradation. The social component shows the smallest performance drop when removed (MAE increases 3.8%, Pearson decreases 6.2%, QWK decreases 7.7%). This might appear counterintuitive given that social communication deficits define autism diagnosis (Guha 2014), but the result reflects an information property of our setting: the current data modalities do not include facial expression, voice, or conversational content, which are primary channels through which social communication deficits manifest clinically. This limits the incremental contribution of the social module beyond what body-level visual features already provide. Nevertheless, the substantial Pearson reduction of 6.2% indicates that the module still adds value by imposing structured and interpretable latent representations.

The motor component demonstrates a more substantial contribution, with removal increasing MAE by 5.4% and reducing Pearson correlation by 10.5%. This finding aligns with emerging evidence that motor impairments are not merely comorbid but core features systematically linked to autism severity (Fournier et al. 2010, MacDonald et al. 2013). While visual features encode movement appearance, they provide limited access to movement dynamics such as smoothness, coordination, and temporal execution. The motor control module addresses this gap by projecting kinematic features into a motor-specific latent space that captures these dynamic properties in a manner complementary to visual information. From a clinical perspective, this result suggests that computational assessment systems should attend to motor patterns even when they are not part of formal diagnostic criteria, as motor behaviors may reflect underlying neurological differences relevant to both motor and social-cognitive function.

Together, the complementary contributions of both components support the multidimensional view of autism that motivates our design. Severity is not explained by a single factor but reflects interacting differences across social and motor domains. The learnable theory weights enable adaptive integration of these domains in a way that reflects individual variability.

### 5.3. Comparison with Alternative Fusion Designs

Ablation studies establish that individual theory components contribute, but do not fully isolate the *fusion mechanism*’s contribution. To address this, we compare against multiple alternatives controlling for model capacity while varying fusion design: (1) **Per Theory**, in which each theory has its own prediction head and the final output is a weighted sum of theory-specific predictions; (2) **Mixture of Experts (MOE)**, where each theory is an expert that produces a prediction and the model combines them via the same learnable theory weights; (3) **Gated**, where theory weights gate the concatenated features before a shared MLP and regression head. Our approach (**weighted sum**) forms a single representation as a weighted sum of theory features and passes it through one regression head, avoiding an extra fusion MLP and remaining fully linear in the theory space. Table 3 summarizes the results.

**Table 3.**
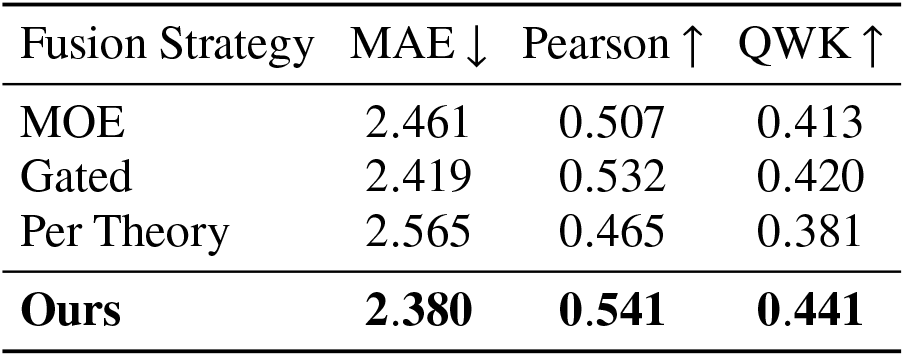
Comparison with Alternative Multimodal Fusion Designs.

Across all three metrics, ours is the most effective: it achieves the lowest MAE (2.380), the highest Pearson correlation (0.541), and the highest QWK (0.441). Relative to the strongest alternative, Gated, our approach reduces MAE from 2.419 to 2.380, increases Pearson from 0.532 to 0.541, and increases QWK from 0.420 to 0.441. Compared to MOE, the gains are larger on agreement-based metrics: QWK improves from 0.413 to 0.441, with MAE improving from 2.461 to 2.380 and Pearson from 0.507 to 0.541. The Per Theory design performs worst overall (MAE 2.565, Pearson 0.465, QWK 0.381), indicating that deferring fusion to the level of independent theory-specific predictions loses information that is useful for accurate severity estimation.

These results support our central design choice: fuse at the *representation level* using an *instance-specific combination* of theory latents, then apply a *single linear regression head*. The alternatives introduce additional nonlinearities that can increase flexibility but also complicate the mapping between theory channels and the final score. In Per Theory and MOE, each theory produces its own prediction, and the final output is a mixture of scalar outputs. This makes the gating weights difficult to interpret as contributions of clinically meaningful features, because each expert can internally re-encode or compensate for missing signal in a theory-specific way before emitting a scalar, and different experts can implement different score-to-feature mappings. The degradation in Pearson (e.g., 0.465 for Per Theory versus 0.541 for ours) is consistent with this: mixing predictions does not reliably preserve a stable, monotone relationship between latent behavioral evidence and the final severity score.

The Gated model is closer in spirit to our approach, but it gates a concatenated feature vector and then passes it through a shared MLP before regression. This architecture weakens interpretability in two ways. First, after gating, the fusion MLP can introduce cross-theory interactions that make the effect of *α*_soc_ and *α*_mot_ non-identifiable: the same output can be achieved via different internal interaction patterns, so the weights no longer correspond to a clean “percent attribution” across constructs. Second, the MLP can re-scale and re-mix gated dimensions, so a larger theory weight does not necessarily imply a larger causal contribution of that theory channel to *ŷ*. Empirically, even with this extra flexibility, Gated remains slightly worse than our linear weighted-sum design on all metrics, suggesting that the additional nonlinearity does not translate into better generalization in this clinical, low-sample setting.

Overall, Table 3 indicates that our fusion choice is not a cosmetic architectural preference. It delivers the best predictive accuracy while preserving a transparent decomposition of severity into theory-aligned components. This is precisely the design objective for clinical modeling: improve generalization while keeping the model’s decision pathway interpretable at the level of symptom constructs.

### 5.4. Impact of Alignment Mask Design

Next, we examine a key design choice within the cross-attention module, namely whether explicitly modeling joint to patch correspondence through an alignment mask improves cross-modal alignment and downstream prediction. Broadly, three approaches can be used to establish correspondence between skeleton joints and image patches. A purely data-driven approach applies no constraints, allowing the model to learn arbitrary attention patterns from data alone. While flexible, this strategy is prone to spurious correlations when training data are limited. A fixed anatomical prior approach predefines which joints attend to which image regions based on body structure, for example restricting hand joints to attend only to hand-region patches. This approach is interpretable but rigid, and it cannot adapt to individual variation or capture unexpected yet informative cross-region dependencies. A third approach introduces a learnable structural bias that initializes correspondences using anatomically plausible priors while allowing them to be refined through gradient-based learning. This strategy balances domain knowledge with data-driven adaptation. Our learnable alignment mask follows this third approach by encoding soft correspondence priors that bias attention toward anatomically plausible alignments while remaining adjustable during training. We compare three configurations: no alignment mask, fixed alignment mask, and learnable alignment mask. Results are shown in Table 4.

**Table 4.**
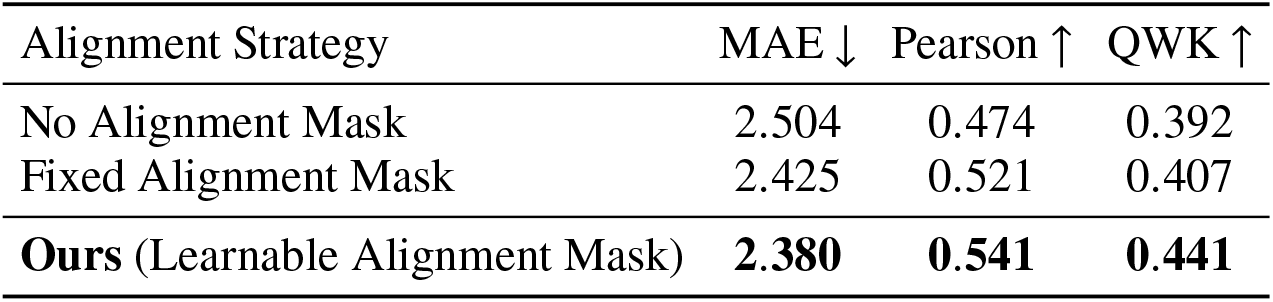
Impact of Alignment Mask Design on Model Performance.

| Alignment Strategy | MAE ↓ | Pearson ↑ | QWK ↑ |
| --- | --- | --- | --- |
| No Alignment Mask | 2.504 | 0.474 | 0.392 |
| Fixed Alignment Mask | 2.425 | 0.521 | 0.407 |
| **Ours (Learnable Alignment Mask)** | **2.380** | **0.541** | **0.441** |

The learnable alignment mask achieves the best performance across all metrics. Compared to the no-mask and fixed-mask variants, MAE is reduced by 1.9% to 5.0%, Pearson correlation is increased by 3.8% to 14.1%, and QWK is increased by 8.4% to 12.5%. Notably, the fixed anatomical mask improves over the no-mask configuration but still underperforms the learnable mask. This pattern can be explained by the construction of SKEPXEL representations, which transform skeleton sequences into pseudo-images using superpixel encoding that prioritizes spatiotemporal compactness rather than strict anatomical alignment. While spatial proximity remains semantically meaningful, the resulting patch layout does not correspond one-to-one with anatomical topology. As a result, fixed priors that assume specific joints always map to specific patches inject some useful structure but also impose rigid correspondences that cannot adapt when the SKEPXEL layout diverges from anatomical topology. In contrast, the learnable mask treats alignment as a hypothesis to be refined rather than a constraint to be enforced, allowing the model to discover semantically meaningful correspondences in the learned feature space through backpropagation.

The no-mask configuration performs worst across all three metrics. This indicates that while attention mechanisms can discover useful correspondences without explicit guidance, they typically require more data and may converge to suboptimal solutions. Introducing an alignment mask provides inductive bias that guides attention toward plausible structures, with the learnable variant further preserving flexibility for task-specific adaptation.

Overall, these results show that domain-knowledge-informed model design can guide learning effectively when implemented in a flexible manner. Hard, non-adaptive priors risk imposing incorrect structure when assumptions do not align with the data, while purely data-driven learning lacks guidance and may converge inefficiently. Learnable structural priors combine the advantages of both approaches by integrating anatomical intuition with data-driven refinement.

### 5.5. Impact of Cross-Attention Direction

The final design choice we examine concerns a boundary condition often under-specified in multimodal attention: the *directionality* of information flow. When integrating two modalities via cross-attention, which should “query” the other? Should visual features query kinematic (image-to-skeleton), kinematic query visual (skeleton-to-image), or both (bidirectional)? Table 5 reports results for these three configurations.

**Table 5.**
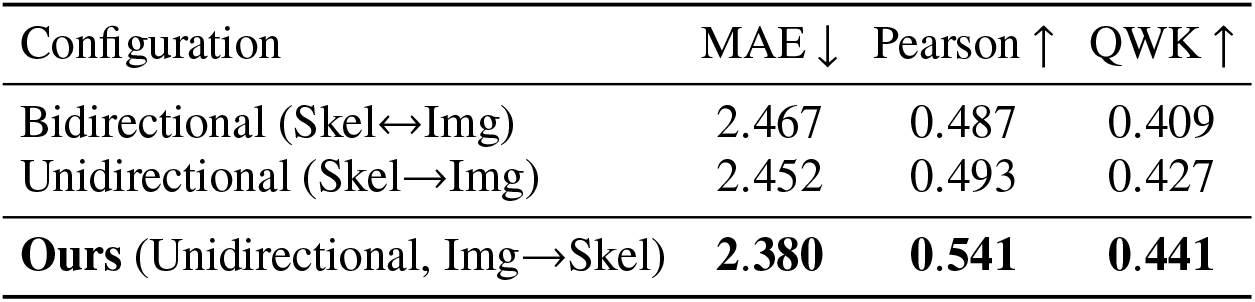
Performance Comparison of Cross-Attention Direction Configurations.

| Configuration | MAE ↓ | Pearson ↑ | QWK ↑ |
| --- | --- | --- | --- |
| Bidirectional (Skel↔Img) | 2.467 | 0.487 | 0.409 |
| Unidirectional (Skel→Img) | 2.452 | 0.493 | 0.427 |
| **Ours** (Unidirectional, Img→Skel) | **2.380** | **0.541** | **0.441** |

The results show a clear ordering. Image-to-skeleton attention achieves the best performance, with MAE of 2.380, Pearson correlation of 0.541, and QWK of 0.441. Skeleton-to-image attention performs second best, with MAE of 2.452, Pearson of 0.493, and QWK of 0.427. Bidirectional attention performs worst, with MAE of 2.467, Pearson of 0.487, and QWK of 0.409. Relative to the bidirectional configuration, image-to-skeleton improves MAE by 3.5%, Pearson correlation by 11.1%, and QWK by 7.8%. In contrast, skeleton-to-image improves performance by only 0.6% in MAE, 1.2% in Pearson, and 4.4% in QWK. These results confirm that attention directionality materially affects performance and that visual-to-kinematic information flow is most effective for autism severity prediction.

Three complementary factors help explain the superiority of image-to-skeleton attention. First, representational richness differs across modalities. Visual features derived from SKEPXEL encode global contextual information such as body posture, spatial relationships, and overall configuration, whereas kinematic features encode local, joint-level motion patterns. When image patches query skeleton joints, attention effectively conditions local kinematic signals on global visual context. This mirrors clinical reasoning, where movement interpretation depends on surrounding posture and scene context. Reversing the direction weakens this asymmetry, as local kinematics provide limited context for interpreting global visual structure. Second, unidirectional attention offers implicit regularization. The image-to-skeleton configuration uses fewer parameters. This reduced complexity is beneficial for small clinical datasets. The worse performance of bidirectional attention suggests that although symmetric information exchange is theoretically appealing, the added complexity does not translate into improved generalization and may instead increase overfitting risk. Third, semantic redundancy arises in the skeleton-to-image configuration. SKEPXEL representations are constructed from skeleton data, producing pseudo-images that encode kinematic information. When skeleton joints query image patches, the model may effectively retrieve transformed versions of information originating from the same source, resulting in circular or redundant information flow. In contrast, image-to-skeleton attention allows ViT-enriched visual features to provide complementary contextual information to MS-G3D-enriched kinematic representations, leading to more informative fusion.

## 6. Conclusion

This paper presents a novel theory-driven deep learning model for automated autism severity assessment from privacy-preserving temporal skeleton data. Instead of formulating severity prediction as a single black-box regression task, we explicitly decompose behavior into two clinically grounded constructs, social communication and motor control, and encode this decomposition into the model architecture. Clinical theory shapes the full pipeline, from representation learning to cross-modal alignment and final fusion, so that predictive accuracy is achieved with an interpretable internal structure. Concretely, the model integrates visual features with kinematic features using unidirectional image-to-skeleton cross-attention, supported by a learnable alignment mask that imposes soft priors over patch–joint correspondences. Construct-level reasoning is then realized through theory-specific processing blocks before latent projection: the Social Attention Block applies self-attention over image patches prior to pooling to capture holistic posture and orientation patterns relevant to social communication, while the Motor Coordination Block explicitly models bilateral structure and left–right asymmetry across joints to emphasize coordination cues relevant to motor control. This construct–modality alignment ensures that each latent channel corresponds to a coherent behavioral dimension rather than an arbitrary feature split. Finally, the construct-aligned latent embeddings are fused using instance-specific, normalized theory weights, producing an interpretable decomposition of each prediction into social and motor contributions. Empirical results show that this theory-guided design achieves state-of-the-art predictive performance while providing clinically grounded explanations that can support clinical decision-making and hypothesis generation.

The theoretical contributions of this work extend prior research on theory-driven algorithm design for healthcare applications. By encoding clinical theories as architectural components, we show how domain knowledge can be operationalized not only as input features or post-hoc interpretations, but as structural elements that shape model learning throughout training. Explicit representation of social communication and motor control as distinct latent channels enables empirical testing of theoretical hypotheses that are inaccessible to atheoretical black-box models. Ablation studies confirm that both constructs contribute measurably to predictive accuracy, providing empirical support for the multidimensional conceptualization of autism motivating contemporary clinical research. Notably, analysis of the learned theory weights reveals systematic relationships between symptom profiles and overall severity. Social communication deficits become increasingly prominent predictors at higher severity levels, whereas motor abnormalities play relatively larger roles at the milder end of the spectrum. This pattern emerges directly from the learned representations and aligns with clinical observations suggesting that motor signatures may be particularly informative biomarkers for individuals with subtler social symptom presentations. Our model therefore functions not only as a predictive model, but also as a tool for theory discovery that enables computational investigation of symptom heterogeneity informing both clinical understanding and intervention design.

The practical implications of this research span clinical assessment, intervention planning, and broader deployment of artificial intelligence in healthcare settings. The achieved performance represents substantial gains in diagnostic accuracy relative to existing methods. Beyond predictive performance, the interpretable theory weights provide clinicians with transparent explanations of model outputs in terms of symptom dimensions already used in clinical assessment. When the model assigns high weight to motor abnormalities for an individual, clinicians can verify this against their own observations, supporting appropriate trust calibration and identifying cases that may warrant closer examination. This transparency addresses a key barrier to clinical AI adoption, namely clinician reluctance to rely on predictions that cannot be understood or verified. In addition, the instance-specific symptom profiles generated by the model offer potential utility for personalized intervention planning. Individuals whose predictions are dominated by motor weights may benefit from motor-focused interventions, while those with elevated social communication weights may respond more strongly to social skills training. Although such applications require validation beyond the scope of this study, our model establishes a foundation for precision approaches that tailor intervention strategies to individual symptom profiles rather than applying uniform treatment protocols.

Several limitations of this work suggest directions for future research. The two-way decomposition into social communication and motor control, while grounded in clinical theory, is not exhaustive. Autism symptomatology also includes sensory processing differences, restricted and repetitive behaviors, language patterns, and other dimensions not explicitly modeled here. Future work could extend our model with additional theory-aligned channels, potentially informed through collaboration with clinical experts. The modest correlations between theory weights and ADOS total scores indicate that although systematic relationships exist, substantial individual variability remains unexplained, reflecting the inherent heterogeneity of autism. Moreover, the current modalities do not include voice, conversational content, or facial expression information, which are central channels through which social communication deficits manifest. Incorporating these modalities may strengthen the social communication component in future iterations. Finally, although our model provides interpretability by design through exposed theory weights, rigorous evaluation with clinical end-users is necessary to determine whether these explanations are perceived as meaningful and actionable in real-world assessment workflows.

## Data Availability

All data produced in the present study are available upon reasonable request to the authors

